# Characteristics of children and antigen test performance at a SARS-CoV-2 community testing site

**DOI:** 10.1101/2021.07.06.21259792

**Authors:** Laura Ford, Melissa J. Whaley, Melisa M. Shah, Phillip P. Salvatore, Hannah E. Segaloff, Augustina Delaney, Dustin W. Currie, Lauren Boyle-Estheimer, Michelle O’Hegarty, Clint N. Morgan, Jennifer Meece, Lynn Ivacic, Natalie J. Thornburg, Azaibi Tamin, Jennifer L. Harcourt, Jennifer M. Folster, Magdalena Medrzycki, Shilpi Jain, Phili Wong, Kimberly Goffard, Douglas Gieryn, Juliana Kahrs, Kimberly Langolf, Tara Zochert, Jacqueline E. Tate, Christopher H. Hsu, Hannah L. Kirking

## Abstract

**Background:** Performance characteristics of SARS-CoV-2 antigen tests among children are limited despite the need for point-of-care testing in school and childcare settings. We describe children seeking SARS-CoV-2 testing at a community site and compare antigen test performance to real-time reverse transcription-polymerase chain reaction (RT-PCR) and viral culture.

**Methods:** Two anterior nasal specimens were self-collected for BinaxNOW antigen and RT-PCR testing, along with demographics, symptoms, and exposure information from individuals ≥5 years at a community testing site. Viral culture was attempted on residual antigen or RT-PCR positive specimens. Demographic and clinical characteristics, and the performance of SARS-CoV-2 antigen tests, were compared among children (<18 years) and adults.

**Results:** About one in ten included specimens were from children (225/2110); 16.4% (37/225) were RT-PCR positive. Cycle threshold values were similar among RT-PCR positive specimens from children and adults (22.5 vs 21.3, p=0.46) and among specimens from symptomatic and asymptomatic children (22.5 vs 23.2, p=0.39). Sensitivity of antigen test compared to RT-PCR was 73.0% (27/37) among specimens from children and 80.8% (240/297) among specimens from adults; among specimens from children, specificity was 100% (188/188), positive and negative predictive value were 100% (27/27) and 94.9% (188/198) respectively. Virus was isolated from 51.4% (19/37) of RT-PCR positive pediatric specimens; all 19 had positive antigen test results.

**Conclusions:** With lower sensitivity relative to RT-PCR, antigen tests may not diagnose all positive COVID-19 cases; however, antigen testing identified children with live SARS-CoV-2 virus.

## INTRODUCTION

Coronavirus disease 2019 (COVID-19) cases in children in the United States reached over two million in December 2020 [1]. While children are more likely to be asymptomatic or have milder illness [2-4], children can transmit the virus to other children and adults [5-8].

SARS-CoV-2 testing to identify and isolate infected individuals and quarantine their close contacts is an important part of COVID-19 prevention efforts [9]. Although diagnostic testing is recommended regardless of age, children with symptoms and/or exposures are less likely to undergo diagnostic testing than adults [3, 10-13]. Therefore, screening of asymptomatic individuals, including children, has been suggested as a prevention strategy [3, 10-13].

Community surge testing and screening programs pair well with antigen-based tests due to their low cost and provision of rapid results without specialized equipment [14-18]. Antigen-based tests detect the presence of a specific viral antigen and may be performed on nasopharyngeal or nasal swab specimens from persons of any age [19]. Antigen tests are most sensitive for detecting specimens with high viral loads and correlate well with viral culture [19, 20]. When testing symptomatic adults, antigen-based tests have high concordance with real-time reverse transcription-polymerase chain reaction (RT-PCR), which is the gold standard test for SARS-CoV-2 detection [14, 19]. Lower sensitivity and specificity have been observed in asymptomatic adults and in adults who undergo testing more than seven days from symptom onset [21, 22].

However, there are limited data on antigen test performance in children, who are more likely to be asymptomatic or mildly symptomatic with SARS-CoV-2 infection. Given rapid and wide distribution of antigen tests for both diagnostic and screening testing in children, there is greater need to understand their performance. In this investigation, we describe children who sought SARS-CoV-2 testing at a community testing site and compare BinaxNOW (Abbott Laboratories, Abbott Park, IL) SARS-CoV-2 antigen test performance in children relative to RT-PCR and viral isolation in culture.

## METHODS

### Investigation participants and enrollment

Beginning on November 4, 2020, a COVID-19 surge community testing site opened in Oshkosh, Wisconsin, and has been offering SARS-CoV-2 BinaxNOW antigen or RT-PCR testing to the public. Individuals could be tested regardless of symptoms or exposures. In collaboration with the Wisconsin Department of Health Services, the University of Wisconsin System, and the U.S. Department of Health and Human Services, the Centers for Disease Control and Prevention (CDC) conducted an investigation at the surge testing site between November 16 and December 15 to evaluate and validate performance of the site-selected BinaxNOW antigen test in a community setting [22]. Individuals ≥5 years of age who sought testing at this site and received an antigen test were eligible to participate in the CDC investigation, and a convenience sample was recruited. For those who chose to participate, patient information was collected, along with self-collected paired nasal swabs for both antigen and RT-PCR testing. This activity was reviewed by CDC and was conducted consistent with applicable federal law and CDC policy.^1^

### Data collection and testing algorithm

Participants completed a self-administered standardized paper questionnaire, which captured demographics, symptoms, and known exposure to COVID-19 case(s) in the 14 days prior to specimen collection. Parents or guardians completed questionnaires for children who were unable to do so on their own. Approximately 30 minutes after community testing site staff completed a participant’s antigen test, the participant provided two additional observed self-collected anterior nasal swabs, sometimes with assistance from a household member (e.g. for young children or persons with disabilities). Participants were instructed to simultaneously insert one swab into each nostril, rotate five times, then switch nostrils and repeat the process. CDC staff performed the antigen test with one of the two additional nasal swabs per manufacturer instructions [20]. The second simultaneously collected nasal swab was placed in transport media (UTM, Remel, Lenexa, KS, US) for RT-PCR testing. COVID-19 TaqPath RT-PCR testing was completed at the Marshfield Clinic Research Institute [23]. Specimens with a cycle threshold (Ct) value (indicating levels of viral RNA) of ≤ 37 for at least two of three SARS-CoV-2 gene targets (ORF1ab, S-gene, and N-gene) on the RT-PCR assay were considered positive.^23^ Virus isolation was attempted using Vero CCL-81 cell suspension in 96-well format from residual RT-PCR specimens from all patients who tested positive by either RT-PCR or antigen testing [24].

An antigen test was indeterminant when a control line did not appear or remained blue [20]. Inconclusive RT-PCR results were only positive for one of the three targets [23]. Participants with indeterminate antigen results or inconclusive RT-PCR results were excluded from analyses.

Some individuals registered for antigen tests on more than one day throughout the investigation period and could therefore contribute multiple specimens to the investigation if tested on different days.

### Analysis

We defined children as participants <18 years old and adults as ≥18 years old. We categorized children in the following age groups: 5 to 8 years old, 9 to 12 years old, 13 to 15 years old, and 16 to 17 years old. Participants who reported ≥1 of COVID-19 symptoms, listed in the Council for State and Territorial Epidemiologists (CSTE) clinical criteria [25], at the time of specimen collection were considered symptomatic; participants who did not report any COVID-19 symptoms at the time of specimen collection were considered asymptomatic. Participants met the CSTE clinical criteria for COVID-19 (a surveillance case definition used by public health surveillance systems within the United States) if they had a cough, shortness of breath, difficulty breathing, a new loss of taste or smell, or had two or more of the following: fever, chills, rigors, muscle aches, headache, sore throat, nausea, diarrhea, fatigue, and congestion [25]. An exposure was defined as reporting being within six feet of a person with a diagnosis of COVID-19 for at least 15 minutes in the past 14 days. We compared demographics, exposures, and symptoms in children to adult participants. We used chi-square tests, and Fisher’s exact tests when cell values were <5, to assess differences among dichotomous/categorical characteristics and considered p-values <0.05 as statistically significant. Median days with interquartile ranges (IQRs) were calculated for the time interval between date of symptom onset and/or last known exposure and the specimen collection date.

Results from the antigen test specimen collected at the same time as the RT-PCR specimen were used for all analyses. With analysis stratified by children and adults, we assessed concordance between antigen and RT-PCR tests using Cohen’s Kappa statistic. Antigen test sensitivity, specificity, positive predicative value (PPV), and negative predicative value (NPV) were calculated with RT-PCR results as the reference; 95% confidence intervals (CI) for each performance characteristic were determined with the exact binomial method. We calculated percent positive by antigen test and RT-PCR for all age categories. To further evaluate test differences, we compared N gene Ct values, detected in all positive RT-PCR specimens, and viral culture results. Test results were compared based on age group, symptom and exposure status. Two-sided Mann-Whitney tests were used for the comparison of Ct values. Statistical analysis was performed in SAS 9.4, SAS Institute Inc. and figures were prepared with R version 4.0.2.

## RESULTS

### Study population

Between November 16 and December 15, 2020, data on 2,127/9,473 specimens tested at the community surge testing site were collected; thirteen specimen pairs with inconclusive or missing RT-PCR results and four with indeterminate antigen test results were excluded from further analysis. Males provided 42.9% (905/2110) and non-Hispanic Whites provided 88.9% (1876/2110) of specimens (Table 1). Among all specimens, 89.3% (1885/2110) were collected from 1,807 adults and 10.7% (225/2110) were collected from 217 children.

**Table 1:**
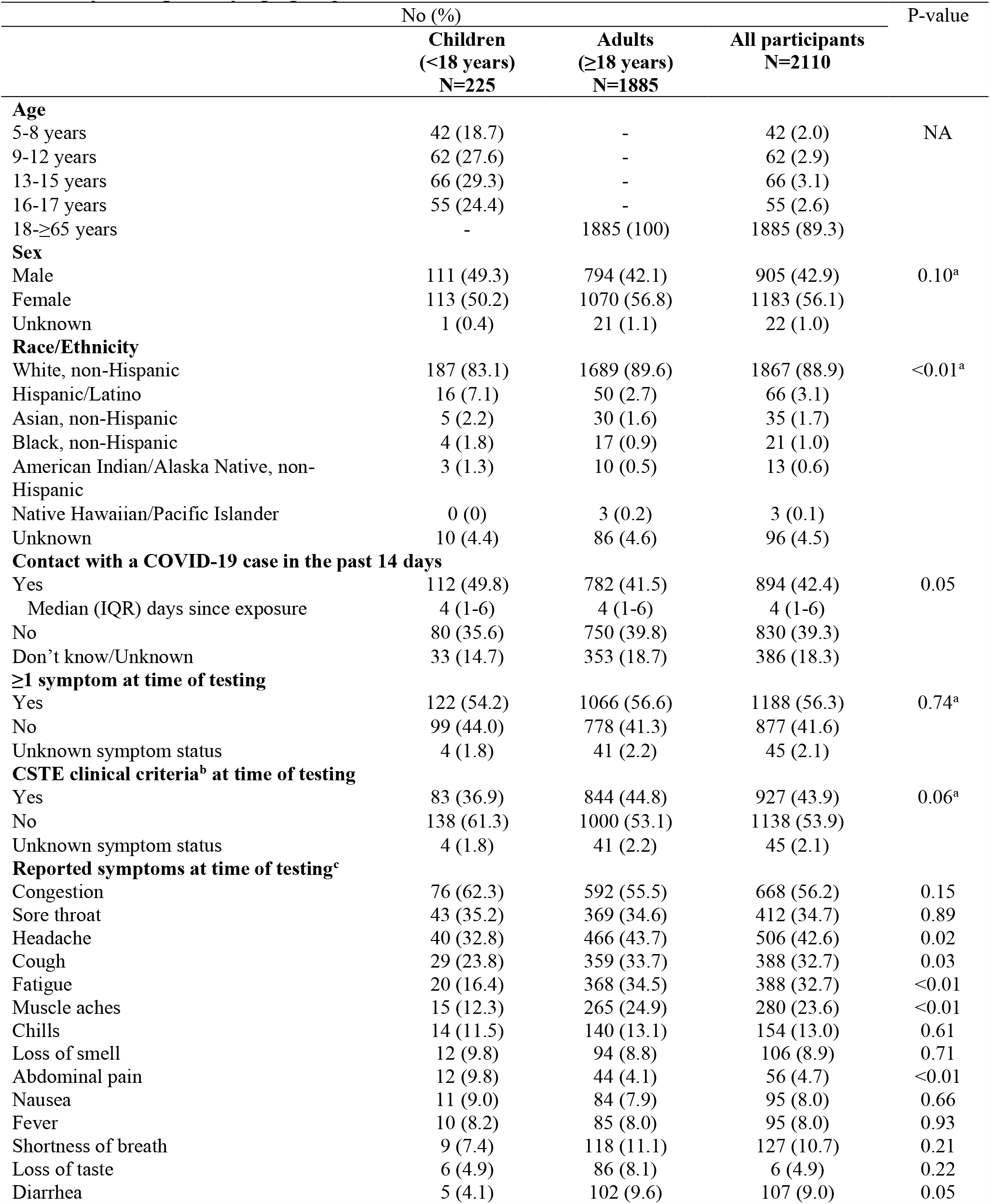
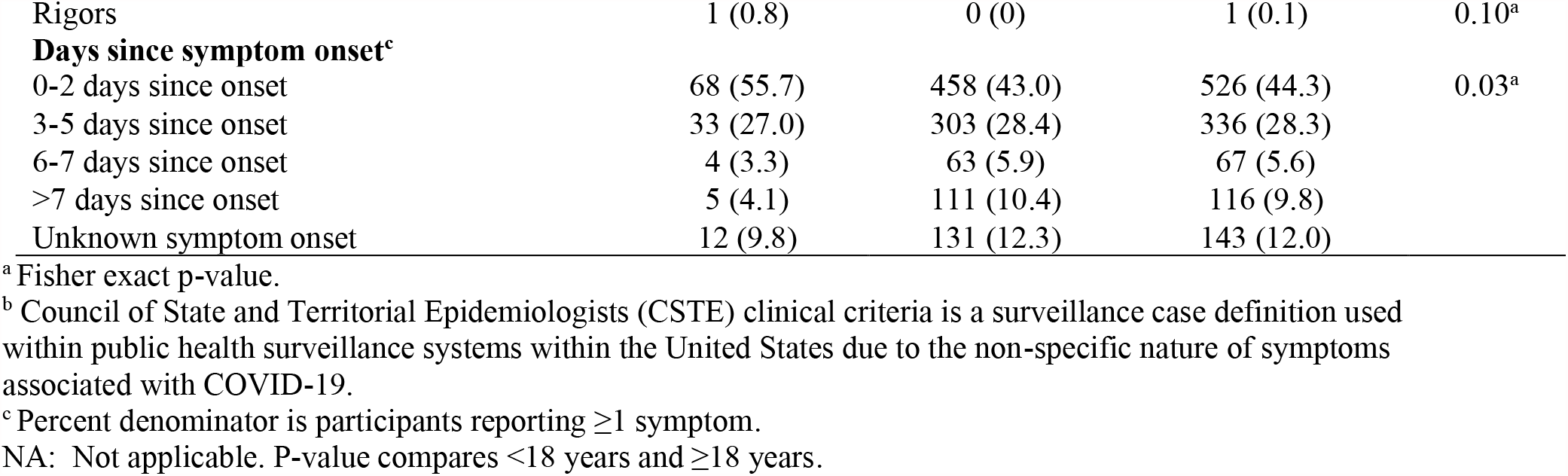
Demographic information, exposure and symptoms of participants testing at a community testing site by age group, Wisconsin, November-December 2020

Among specimens from children, 49.3% (111/225) were from males and 83.1% (187/225) were from non-Hispanic Whites. In this population, 18.7% (42/225) were from participants aged 5-8 years, 27.6% (62/225) were from participants aged 9-12 years, 29.3% (66/225) were from participants aged 13-15 years, and 24.4% (55/225) were from participants aged 16-17 years.

Comparing specimens from adults and children, 41.5% (782/1885) were from adults who reported exposure to a COVID-19 case in the past 14 days, versus 49.8% (112/225) of specimens from children (p=0.05). Exposures among children aged 16-17 years were significantly higher than adults (60%, 33/55, p=0.02, Supplementary Table 1). Most specimens were from symptomatic participants for both children (54.2%; 122/225) and adults (56.6%; 1066/1885) (p=0.74). Compared to adults, a higher proportion of specimens were from symptomatic children who reported only one symptom (35.2% vs 24.1%, p<0.01). Eighty-three (36.9%) child and 844 (44.8%) adult specimens were from individuals reporting symptoms that matched the CSTE clinical criteria for COVID-19 (p=0.06). Specimens were collected a median of two days post-symptom onset from children (IQR 1-3; 9.8% missing onset date) and three days post-symptom onset from adults (IQR 1-5; 12.3% missing onset date).

### RT-PCR positive participants

RT-PCR positivity was 15.8% (297/1885) among specimens from adults and 16.4% (37/225) among specimens from children (Figure 1). Among specimens from children by age group, RT-PCR positivity was 16.7% (7/42) for children 5-8 years, 14.5% (9/62) for children 9-12 years, 9.1% (6/66) for children 13-15 years, and 27.3% (15/55) for children 16-17 years (Figure 1 and Supplementary Figure 1). Specimens from participants aged 16-17 years had significantly higher positivity by RT-PCR than participants aged <16 years (p=0.01) and adults (p=0.02).

**Figure 1:**
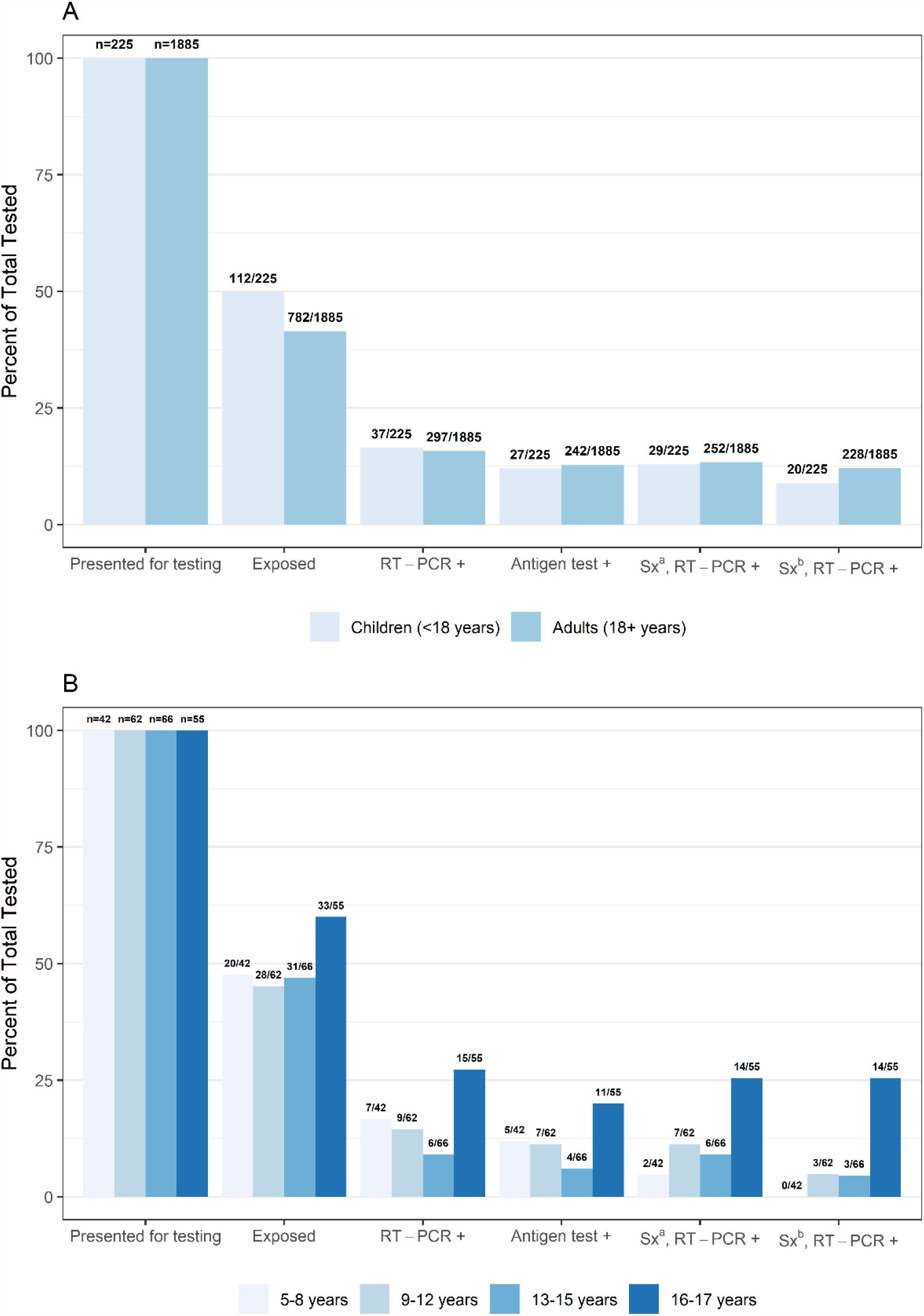
(A) Percent presenting for testing, exposed, real-time reverse transcription-polymerase chain reaction (RT-PCR) or antigen test positive, and (B) positive and symptomatic by age group, collected at a community testing site – Oshkosh, Wisconsin, November-December 2020 ^a^ Sx: Symptomatic defined as reporting ≥1 symptom at specimen collection ^b^ Sx: Symptomatic defined as reporting symptoms meeting the Council of State and Territorial Epidemiologists (CSTE) clinical criteria for COVID-19

The proportion of RT-PCR positive specimens from exposed children (56.8%) and symptomatic children (78.4%) was similar to the proportion of RT-PCR positive specimens from exposed (54.9% p=0.98) and symptomatic (84.8%, p=0.33) adults (Supplementary Tables 2-3). Among RT-PCR positive specimens, a lower proportion were from children who reported symptoms meeting the CSTE clinical criteria than from adults who reported meeting the CSTE clinical criteria (54.1% vs 76.8%, p=0.01).

Median Ct values did not differ between specimens from children and adults (p=0.46, Table 2) or among pediatric age groups (5-8 years: 22.2, 9-12 years: 22.8, 13-15 years: 23.4, and 16-17 years 20.6, p=0.90; Supplementary Table 4). The median Ct value of RT-PCR positive specimens from children did not differ by symptom status or by symptom duration; among RT-PCR positive specimens from adults, the median Ct value also did not differ by symptom status but was lower among specimens from adults tested within seven days of symptom onset compared to adults tested >7 days since symptom onset (Ct value 20.8 vs. 27.7, p<0.01).

**Table 2:**
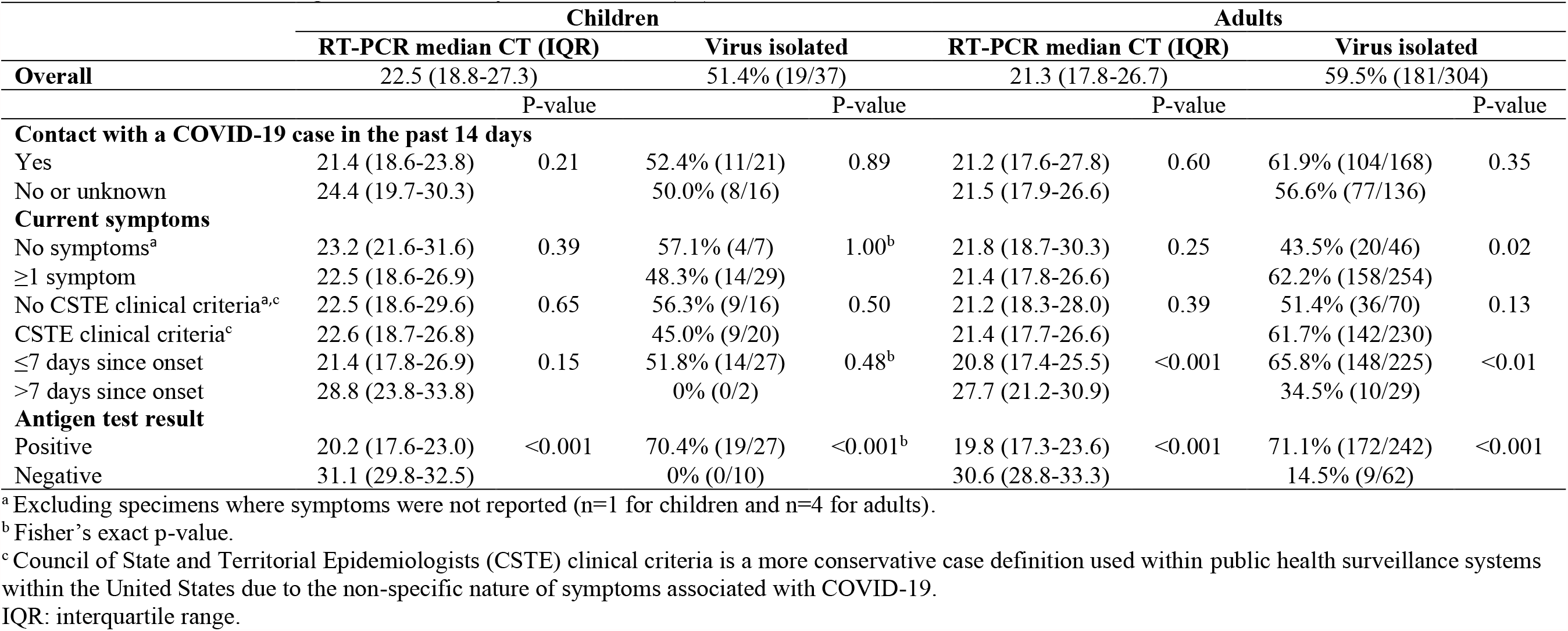
SARS-CoV-2 N-gene RT-PCR cycle threshold (Ct) values and virus isolation, children and adults

Virus was isolated from 51.4% (19/37) of RT-PCR positive specimens from children and 59.5% (181/304) of RT-PCR or antigen positive specimens from adults (Table 2). The proportion of positive specimens with isolated virus did not differ among children by exposure (52.4% exposed, p=0.89) or symptom (48.7% symptomatic, p=1.00) status. However, the proportion of positive specimens with isolated virus differed among adults by symptom status (asymptomatic, 43.5% vs symptomatic, 62.2%, p=0.02) and by symptom duration prior to collection (≤7 days, 65.8% vs >7 days, 34.5%; p<0.01).

### Antigen test performance compared to RT-PCR test and viral isolation

Antigen test positivity was 12.8% (242/1885) among adults and 12.0% (27/225) among children (Figure 1; positivity by pediatric age group in Supplementary Table 3). High concordance between antigen test and RT-PCR test was observed, but lower when testing specimens from children (k 0.82, 95% CI 0.71-0.93) than adults (k 0.87, 95% CI 0.84-0.93). Antigen test sensitivity was 73.0% (27/37, 95% CI 55.9%-86.2%) among specimens from children and 80.8% (240/297, 95% CI 75.9%-85.1%) among specimens from adults; specificity was 100% (188/188, 95% CI 98.1%-100%) and 99.9% (1586/1588, 95% CI 99.5%-100%), respectively; PPV was 100% (27/27, 95% CI 87.2%-100%) and 99.2% (240/242, 95% CI 97.0%-99.9%), respectively; NPV was 94.9% (188/198, 95% CI 90.9%-97.6%) and 96.5% (1586/1643, 95% CI 95.5%-97.4%), respectively (Figure 2 and Supplementary Tables 5-7).

**Figure 2:**
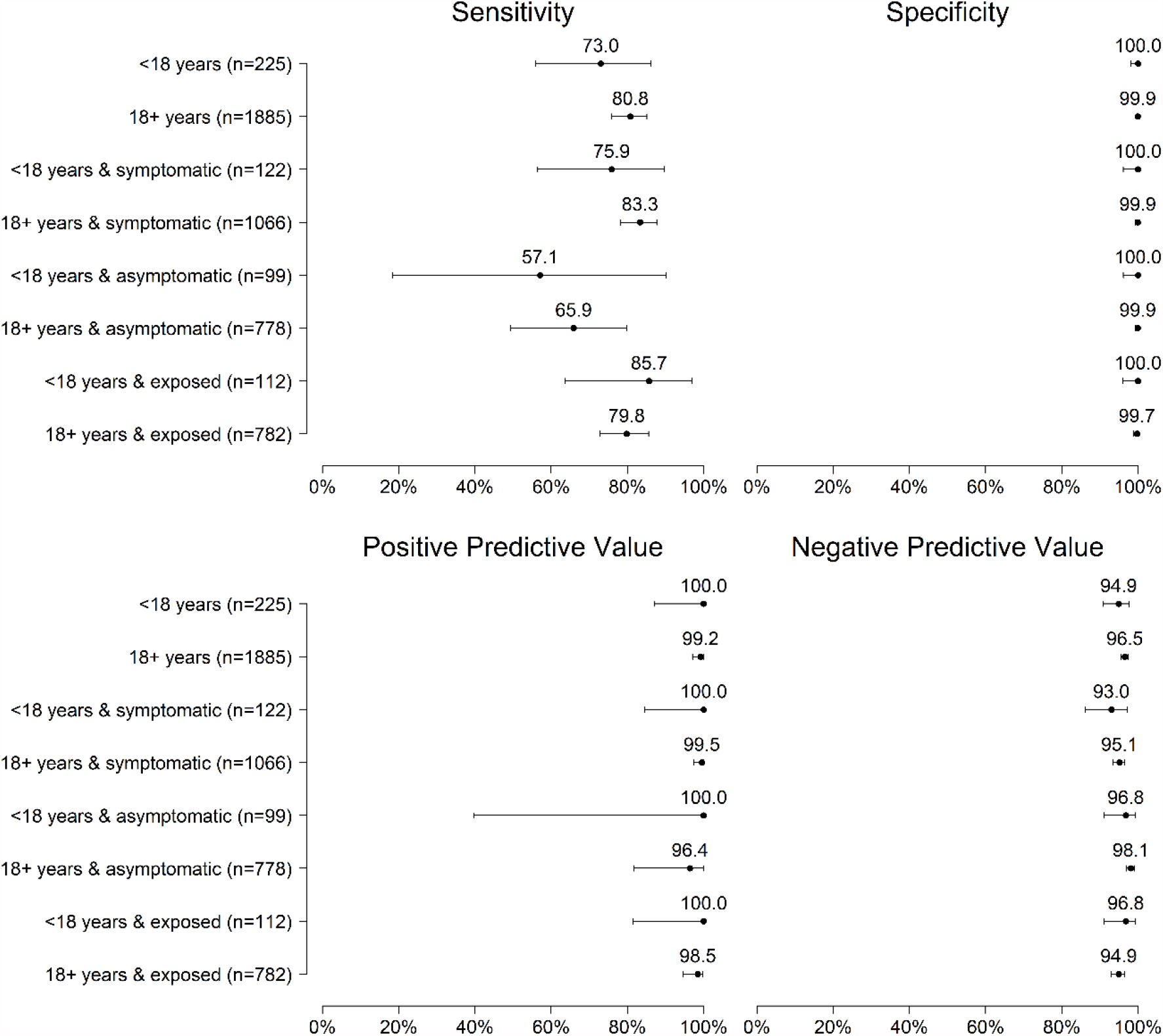
Sensitivity, specificity, positive predictive value, and negative predictive value of antigen test compared with real-time reverse transcription–polymerase chain reaction (RT-PCR) test among pediatric and adult participants overall and by symptom and exposure status, Oshkosh, Wisconsin, November–December 2020

Test sensitivity was 75.9% (95% CI 56.5%-89.7%) among specimens from symptomatic children compared with 57.1% (95% CI 18.4%-90.1%) among asymptomatic children, and 85.7% (6/7) of antigen negative, RT-PCR positive specimens were from symptomatic children tested ≤7 days from symptom onset. Among specimens from children with reported exposure, test sensitivity was 85.7% overall (18/21, 95% CI 63.7%-97.0%), 88.2% with symptoms (15/17, 95% CI 63.6%-98.5%), and 66.7% without symptoms (2/3, 95% CI 9.4%-99.2%) (Supplementary Figure 2). Specificity and PPV were 100% among specimens from symptomatic, asymptomatic, and exposed children. Antigen positive, RT-PCR positive specimens from children were collected a median of four days (IQR 0-6) since last known exposure, while antigen negative, RT-PCR positive specimens were collected a median of two days (IQR 1-6) from known exposure.

Among specimens from both children and adults with a positive RT-PCR result, Ct values for specimens with positive antigen tests were lower than for specimens with negative antigen tests (Table 2, Figure 3, and Supplementary Figure 3). Among RT-PCR and antigen positive specimens from children, virus was isolated from 70.4% (19/27); 73.7% (14/19) were from symptomatic children, 21.1% (4/19) were from asymptomatic children, and 5.3% (1/19) were from children with unknown symptom status. No (0/10) virus was isolated from RT-PCR positive, antigen negative specimens from children.

**Figure 3:**
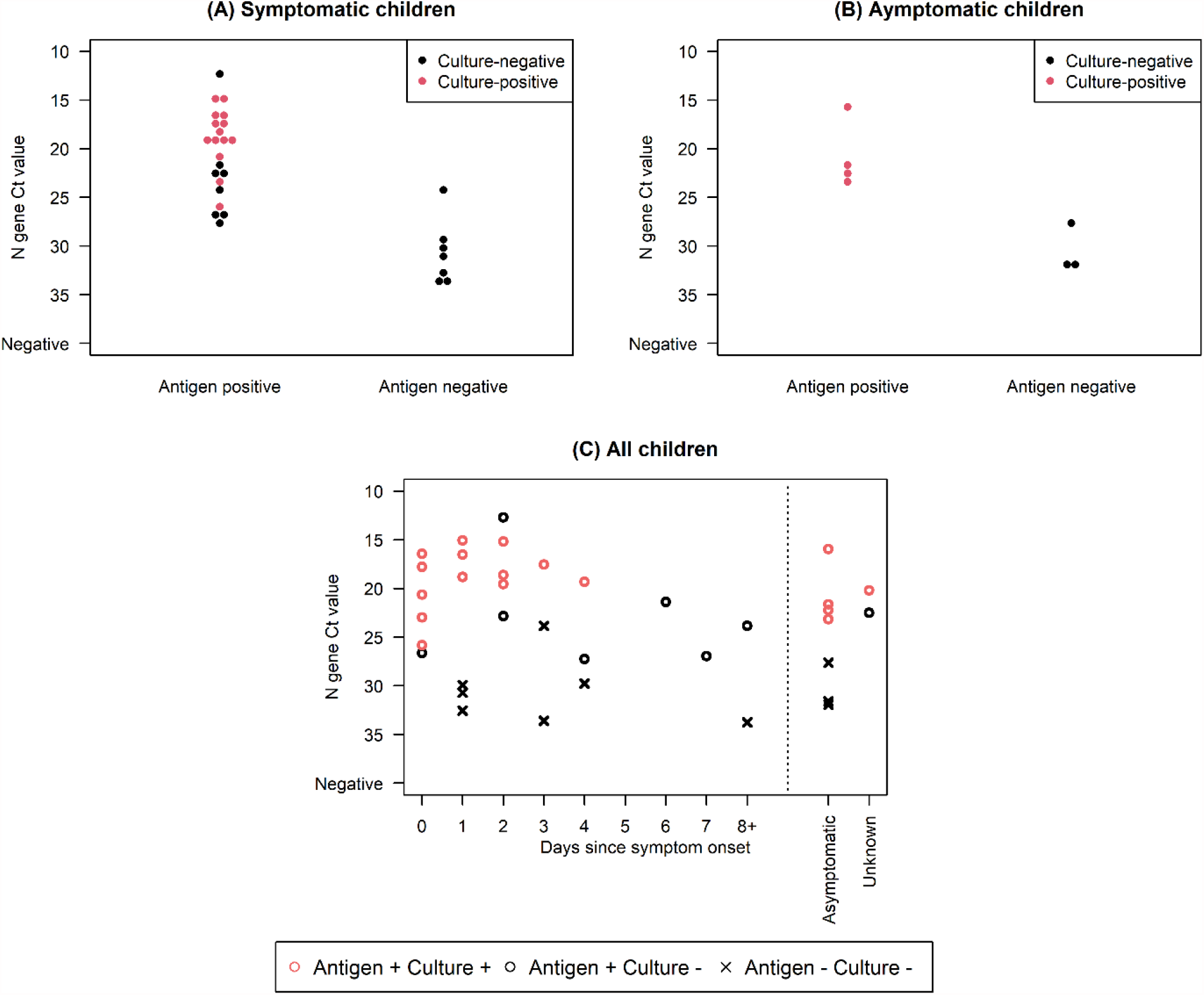
N-gene cycle threshold value distribution and viral isolation among real-time reverse transcription–polymerase chain reaction (RT-PCR) or antigen positive (A) symptomatic children (B) asymptomatic children, (C) all children by days since symptom onset, Oshkosh, Wisconsin, November–December 2020. A & B are excluding 1 child and 4 adults with unknown symptom status

## DISCUSSION

We describe children who sought testing at a community site and provide an opportunity to better understand antigen test performance in children. As of January 2021, 11% of lab-confirmed COVID-19 cases in the US were <18 years of age [26], and in this investigation approximately 11% of specimens and 11% of RT-PCR positive specimens were from children. Over half of pediatric specimens tested were from symptomatic children (a similar proportion to adults), but children generally had fewer symptoms. In particular, RT-PCR positive specimens from younger children (5-8 years) were from either asymptomatic children or children only reporting nasal congestion. Although children are often asymptomatic or have mild, non-specific symptoms [2-4], they still can transmit SARS-CoV-2 [4-8]. Here, virus was isolated in 51% of RT-PCR positive specimens from children, and Ct values were similar to adults. While antigen testing sensitivity in specimens from children was 73%, compared to 81% in specimens from adults, positive antigen results were received for all RT-PCR positive specimens from children with isolated virus.

Antigen test sensitivity among children and adults was consistent with similar studies conducted at community or pediatric clinic testing sites [27, 28]. Among specimens from children, antigen test sensitivity was highest (86%) for those with a known exposure, whereby the probability of infection is higher [9, 13]. More older teenagers (16-17 years) reported a known exposure and had a higher percent positivity than adults. However, whereas few young children (5-8 years) were symptomatic almost all RT-PCR positive teenagers 16-17 years were symptomatic, and the percent of specimens from individuals meeting the CSTE clinical criteria appeared to increase by age. Antigen testing may be useful in this high-prevalence and exposed population, particularly if used as part of a serial testing strategy [29]. Testing pediatric populations, particularly teenagers, with any symptoms or possible exposures is important due to their high levels of exposure and risk of community transmission. Confirmatory nucleic acid amplification testing is recommended for a negative antigen result in individuals with symptoms or known exposures [19].

Median Ct values, which indicate levels of viral RNA, were similar by age, in line with other studies [30, 31]. Median Ct values were also similar among specimens from symptomatic and asymptomatic children, which contrasts with previous studies that found lower Ct values in symptomatic compared with asymptomatic individuals, including those who have recovered from infection [14, 18, 32, 33]. While lower Ct values may suggest higher levels of virus, Ct values are not necessarily a measurement of viral loads [14, 18, 32]. Considering the short duration between testing and contact with a COVID-19 case (i.e. 2-4 days) in this investigation, asymptomatic participants may predominantly be pre-symptomatic instead of at the recovery stage. This may explain why we did not observe differences in Ct values by symptom status.

As reported in other studies [18, 20, 27], Ct values were significantly higher among specimens with antigen negative results than those with antigen positive results. In children, antigen testing also performed better with RT-PCR positive specimens that were culture-positive, than with those that were culture-negative. Viral culture may be insensitive and viral isolation may not perfectly correlate with infectiousness [14, 18, 32]; but it is noteworthy that we detected no live virus in specimens from children with negative antigen results.

Antigen tests were highly specific regardless of symptom status or exposures. There were no antigen positive, RT-PCR negative results in specimens from children, resulting in a PPV of 100% in this moderate-high prevalence setting. Our finding is similar to what was reported at a community testing site in San Francisco and at an outpatient clinic in West Bend, where no antigen positive RT-PCR negative results were received among participants <18 years [16, 28]. The high specificity and ability of these tests to identify those without disease promotes efficient use of scarce public health resources for disease investigation and contact tracing; the specificity also prevents individuals with low-pretest probability from having to isolate unnecessarily due to false positive results. As widespread antigen testing in K-12 schools is considered [9], the advantages and limitations of antigen tests should be taken into account when designing testing strategies.

Our investigation was subject to several limitations. Investigation participants were a convenience sample of largely non-Hispanic White participants and the findings might not be generalizable to other settings. The sample size may have affected the ability to detect significant differences. Furthermore, exposures and symptoms were self-reported, so they may not be accurate or may be symptoms of other respiratory viral infections. Similarly, not many children had a symptom onset >7 days prior to testing, and we were unable to draw conclusions on test performance in this group. Finally, we limited our antigen testing to the BinaxNOW antigen platform, so it is unclear how these results may be generalizable to other antigen platforms.

In conclusion, while children reported fewer symptoms than adults, RT-PCR Ct values and virus isolation results were similar to adults, further supporting that children play a role in transmission [5, 30, 34-36]. Antigen testing was highly specific; estimates suggest that test sensitivity may be highest among exposed children and could be useful in this population regardless of where testing may occur. From this study and others, antigen tests had lower, although not necessarily statistically significant, sensitivity among children compared with adults; this lower sensitivity should be considered when developing diagnostic testing programs.

However, all culture-positive specimens from children had a positive antigen test, indicating that antigen testing identified children with live SARS-CoV-2 virus.

## Data Availability

The datasets generated during and analyzed during the current study are available from the corresponding author on reasonable request.

## ACKNOWLEDGMENTS

The authors would like to thank the contributions of Allen Bateman, Alana Sterkel, and Mary Wedig from the Wisconsin State Laboratory of Hygiene and the members of the Centers for Disease Control and Prevention’s Coronavirus Disease 2019 (COVID-19) Response Team: John Paul Bigouette, Juliana DaSilva, Alicia Fry, Aron Hall, Emiko Kamitane, Sandor Karpathy, Marie Killerby, Nancy Knight, Shirley Lecher, Kaitlin Mitchell, Tarah Somers, and Miriam Van Dyke.

## Footnotes

1 See e.g., 45 C.F.R. part 46.102(l)(2), 21 C.F.R. part 56; 42 U.S.C. §241(d); 5 U.S.C. §552a; 44 U.S.C. §3501 et seq.

