## Supplementary Table for "Characteristics of children and antigen test performance at a SARS-CoV-2 community testing site"

### Supplementary Tables and Figures

**Supplementary Figure 1**: Percent positivity of real–time reverse transcription–polymerase chain reaction (RT–PCR) and antigen tests by symptom status and age group, collected at a community testing site – Oshkosh, Wisconsin, November–December 2020

**Supplementary Figure 2**: Sensitivity, specificity, positive predictive value, and negative predictive value of BinaxNOW antigen test compared with real–time reverse transcription–polymerase chain reaction (RT–PCR) among exposed child and adult participants by symptom status, Oshkosh, Wisconsin, November–December 2020

**Supplementary Figure 3**: N gene cycle threshold value distribution among real–time reverse transcription–polymerase chain reaction (RT–PCR) positive children and adults by symptom status and antigen test result, Oshkosh, Wisconsin, November–December 2020

**Supplementary Table 1**: Exposures and symptoms of children testing at a community testing site by age group, Wisconsin, November–December 2020

**Supplementary Table 2**: Demographic information, exposure, and symptoms of BinaxNOW antigen test or real–time reverse transcription–polymerase chain reaction (RT–PCR) positive participants aged <18 years compared to participants aged ≥18 years, Wisconsin, November–December 2020

**Supplementary Table 3**: Demographic information, exposure, and symptoms of BinaxNOW antigen test or real–time reverse transcription–polymerase chain reaction (RT–PCR) positive participants aged <18 years by age group, Wisconsin, November–December 2020

**Supplementary Table 4**: N–gene RT–PCR mean Ct values comparison by age, Oshkosh, Wisconsin, November–December 2020

**Supplementary Table 5**: Sensitivity, specificity, positive predictive value, and negative predictive value of BinaxNOW antigen test compared with real–time reverse transcription–polymerase chain reaction (RT–PCR) among child and adult participants overall, by symptom status, and by exposure status, Oshkosh, Wisconsin, November–December 2020

**Supplementary Table 6**: Sensitivity, specificity, positive predictive value, and negative predictive value of initial BinaxNOW antigen test compared with real–time reverse transcription–polymerase chain reaction (RT–PCR) among children by age group, overall and by symptom status, Oshkosh, Wisconsin, November–December 2020

**Supplementary Table 7:** Sensitivity, specificity, positive predictive value, and negative predictive value of BinaxNOW antigen test compared with real–time reverse transcription–polymerase chain reaction (RT–PCR) among unique[a] children (n=217), Oshkosh, Wisconsin, November–December 2020

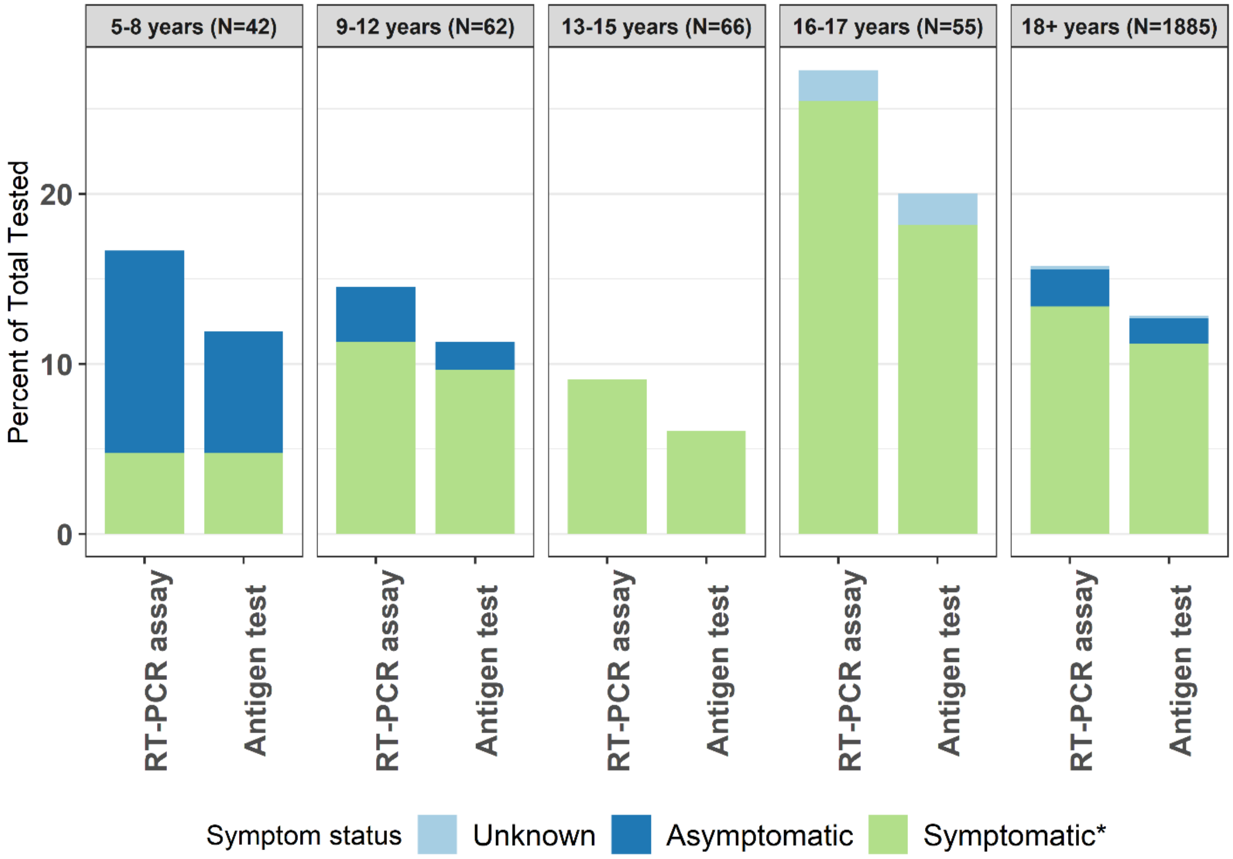


*Symptomatic defined as reporting ≥1 symptom at time of specimen collection

**Supplementary Figure 1**: Percent positivity of real–time reverse transcription–polymerase chain reaction (RT–PCR) and antigen tests by symptom status and age group, collected at a community testing site – Oshkosh, Wisconsin, November–December 2020

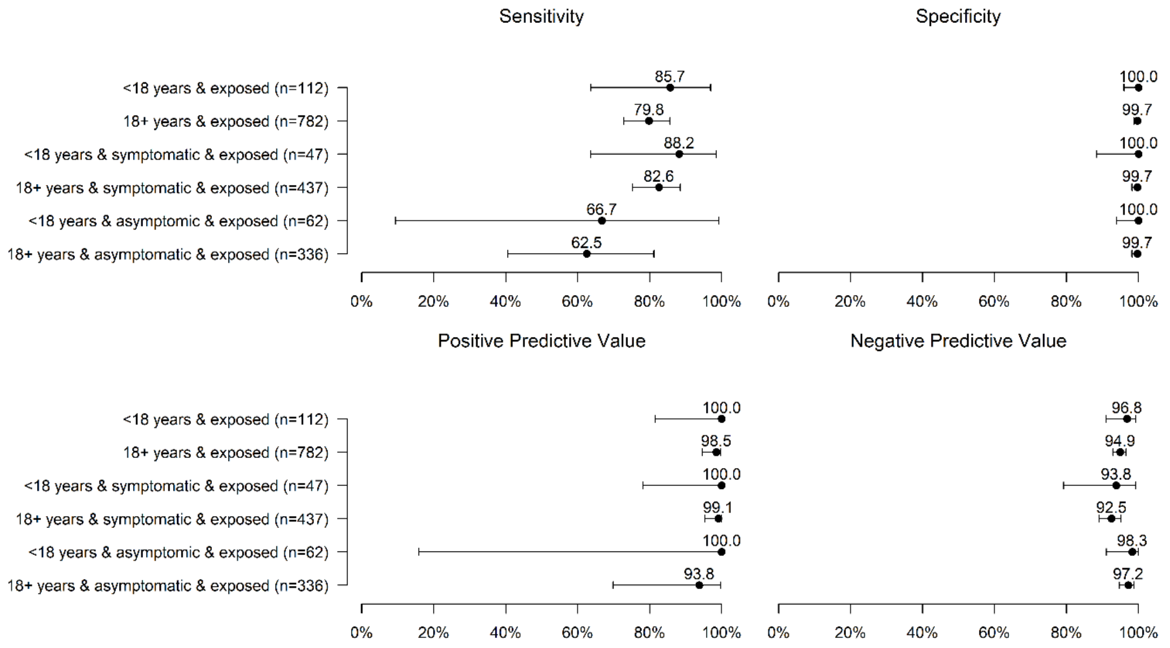


**Supplementary Figure 2:** Sensitivity, specificity, positive predictive value, and negative predictive value of BinaxNOW antigen test compared with real–time reverse transcription–polymerase chain reaction (RT–PCR) among exposed child and adult participants by symptom status, Oshkosh, Wisconsin, November–December 2020

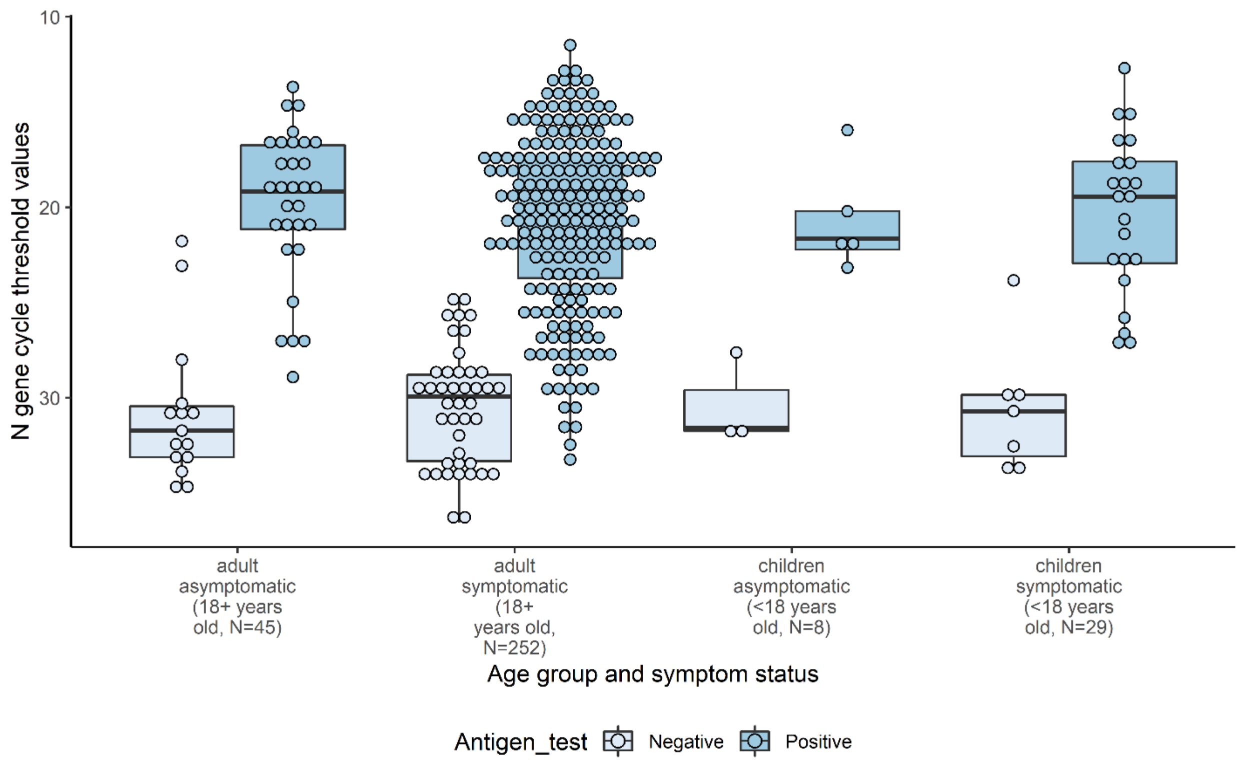


**Supplementary Figure 3**: N gene cycle threshold value distribution among real–time reverse transcription–polymerase chain reaction (RT–PCR) positive children and adults by symptom status and antigen test result, Oshkosh, Wisconsin, November–December 2020

Distribution of N gene Ct values among RT–PCR positive participants stratified by age and symptom status. Light blue circles represent antigen negative results. Dark blue circles represent antigen positive results.

**Supplementary Table 1:** Reported exposures and symptoms of pediatric participants testing at a community testing site by age group, Wisconsin, November–December 2020

| | No (%) | | | |
|---|---|---|---|---|
| | **5–8 years N=42** | **9–12 years N=62** | **13–15 years N=66** | **16–17 years N=55** |
| **Contact with a COVID–19 case in the past 14 days** | | | | |
| Yes | 20 (47.6) | 28 (45.2) | 31 (47.0) | 33 (60.0) |
| No | 16 (38.1) | 27 (43.5) | 22 (33.3) | 15 (27.3) |
| Don't know/Unknown | 6 (14.3) | 7 (11.3) | 13 (19.7) | 7 (12.7) |
| **≥1 symptom at time of testing** | | | | |
| Yes | 24 (57.1) | 33 (53.2) | 35 (53.0) | 30 (54.5) |
| No | 18 (42.9) | 29 (46.8) | 29 (43.9) | 23 (41.8) |
| Unknown symptom status | 0 (0) | 0 (0) | 2 (3.0) | 2 (3.6) |
| **CSTE clinical criteria* at time of testing** | | | | |
| Yes | 12 (28.6) | 15 (24.2) | 28 (42.2) | 26 (47.3) |
| No | 30 (71.4) | 45 (72.6) | 36 (54.5) | 27 (49.1) |
| Unknown symptom status | 0 (0) | 0 (0) | 2 (3.0) | 2 (3.6) |
| **Reported symptoms at time of testing[b]** | | | | |
| Congestion | 19 (79.2) | 17 (51.5) | 19 (54.3) | 21 (70.0) |
| Sore throat | 4 (16.7) | 11 (33.3) | 13 (37.1) | 15 (50.0) |
| Headache | 5 (20.8) | 7 (21.2) | 17 (48.6) | 11 (36.7) |
| Cough | 5 (20.8) | 8 (24.2) | 6 (17.1) | 10 (33.3) |
| Fatigue | 2 (8.3) | 3 (9.1) | 9 (25.7) | 6 (20.0) |
| Muscle aches | 0 (0) | 3 (9.1) | 8 (22.9) | 4 (13.3) |
| Chills | 1 (4.2) | 2 (6.1) | 9 (25.7) | 2 (6.7) |
| Loss of smell | 0 (0) | 1 (3.0) | 6 (17.1) | 5 (16.7) |
| Abdominal pain | 3 (12.5) | 2 (6.1) | 6 (17.1) | 1 (3.3) |
| Nausea | 2 (8.3) | 3 (9.1) | 4 (11.4) | 2 (6.7) |
| Fever | 5 (20.8) | 0 (0) | 3 (8.6) | 2 (6.7) |
| Shortness of breath | 1 (4.2) | 1 (3.0) | 4 (11.4) | 3 (10.0) |
| Loss of taste | 0 (0) | 0 (0) | 5 (14.3) | 1 (3.3) |
| Diarrhea | 1 (4.2) | 1 (3.0) | 0 (0) | 3 (10.0) |
| Rigors | 0 (0) | 0 (0) | 1 (2.9) | 0 (0) |
| **Days since symptom onset[b]** | | | | |
| 0–2 days since onset | 13 (54.2) | 17 (51.5) | 19 (54.3) | 19 (63.3) |
| 3–5 days since onset | 6 (25.0) | 10 (30.3) | 8 (22.9) | 9 (30.0) |
| 6–7 days since onset | 2 (8.3) | 1 (3.0) | 1 (2.9) | 0 (0) |
| >7 days since onset | 1 (4.2) | 1 (3.0) | 3 (8.6) | 0 (0) |
| Unknown symptom onset | 2 (8.3) | 4 (12.1) | 4 (11.4) | 2 (6.7) |

[a] Council of State and Territorial Epidemiologists (CSTE) clinical criteria is a surveillance case definition used within public health surveillance systems within the United States due to the non–specific nature of symptoms associated with COVID–19

[b] Percent denominator is participants reporting ≥1 symptom

**Supplementary Table 2:** Demographic information, exposure and symptoms of BinaxNOW or real–time reverse transcription–polymerase chain reaction (RT–PCR) positive participants aged <18 years compared to participants aged ≥18 years, Wisconsin, November–December 2020

| | **RT–PCR positive no (%)** | | | |
|---|---|---|---|---|
| | **<18 years N=37** | | **≥18 years N=297** | |
| | **Antigen+ N=27** | **Antigen– N=10** | **Antigen+ N=240** | **Antigen– N=57** |
| **Sex** | | | | |
| Male | 14 (51.9) | 5 (50.0) | 109 (45.4) | 26 (45.6) |
| Female | 13 (48.1) | 5 (50.0) | 129 (53.8) | 31 (54.4) |
| Unknown | 0 (0) | 0 (0) | 2 (0.8) | 0 (0) |
| **Race/Ethnicity** | | | | |
| White, non–Hispanic | 20 (74.1) | 10 (100) | 220 (91.7) | 49 (86.0) |
| Hispanic/Latino | 6 (22.2) | 0 (0) | 7 (2.9) | 1 (1.8) |
| Asian, non–Hispanic | 0 (0) | 0 (0) | 4 (1.7) | 1 (1.8) |
| Black, non–Hispanic | 0 (0) | 0 (0) | 0 (0) | 1 (1.8) |
| American Indian/Alaska Native, non–Hispanic | 0 (0) | 0 (0) | 0 (0) | 1 (1.8) |
| Native Hawaiian/Pacific Islander | 0 (0) | 0 (0) | 0 (0) | 0 (0) |
| Unknown | 1 (3.7) | 0 (0) | 9 (3.8) | 4 (7.0) |
| **Contact with a COVID–19 case in the past 14 days** | | | | |
| Yes | 18 (66.7) | 3 (30.0) | 130 (54.2) | 33 (57.9) |
| Median (range) days since exposure | 4 (0–6) | 2 (1–6) | 3 (0–6) | 2 (0–4) |
| No | 5 (18.5) | 4 (40.0) | 59 (24.6) | 16 (28.1) |
| Don't know/Unknown | 4 (14.8) | 3 (30.0) | 51 (21.2) | 8 (14.0) |
| **≥1 symptom at time of testing** | | | | |
| Yes | 22 (81.5) | 7 (70.0) | 210 (87.5) | 42 (73.7) |
| No | 4 (14.8) | 3 (30.0) | 27 (11.2) | 14 (24.6) |
| Unknown symptom status | 1 (3.7) | 0 (0) | 3 (1.3) | 1 (1.8) |
| **CSTE clinical criteria* at time of testing** | | | | |
| Yes | 15 (55.6) | 5 (50.0) | 190 (79.2) | 38 (66.7) |
| No | 11 (40.7) | 5 (50.0) | 47 (19.6) | 18 (31.6) |
| Unknown symptom status | 1 (3.7) | 0 (0) | 3 (1.3) | 1 (1.8) |
| **Reported symptoms at time of testing[b]** | | | | |
| Congestion | 14 (63.6) | 5 (71.4) | 127 (60.5) | 20 (47.6) |
| Sore throat | 9 (40.9) | 1 (14.3) | 61 (29.0) | 10 (23.8) |
| Headache | 7 (31.8) | 1 (14.3) | 88 (41.9) | 13 (31.0) |
| Cough | 5 (22.7) | 2 (28.6) | 103 (49.0) | 17 (40.5) |
| Fatigue | 4 (18.8) | 2 (28.6) | 71 (33.8) | 16 (38.1) |
| Muscle aches | 4 (18.2) | 0 (0) | 71 (33.8) | 9 (21.4) |
| Chills | 1 (4.5) | 0 (0) | 39 (18.6) | 8 (19.0) |
| Loss of smell | 4 (18.2) | 3 (42.9) | 56 (26.7) | 12 (28.6) |
| Abdominal pain | 1 (4.5) | 0 (0) | 7 (3.3) | 1 (2.4) |
| Nausea | 1 (4.5) | 0 (0) | 14 (6.7) | 6 (14.3) |
| Fever | 2 (9.1) | 0 (0) | 38 (18.1) | 2 (4.8) |
| Shortness of breath | 3 (13.6) | 0 (0) | 19 (9.0) | 5 (11.9) |
| Loss of taste | 2 (9.1) | 1 (14.3) | 44 (21.0) | 9 (21.4) |
| Diarrhea | 1 (4.5) | 1 (14.3) | 14 (6.7) | 1 (2.4) |

| | RT–PCR positive no (%) | | | |
|---|---|---|---|---|
| | **<18 years N=37** | | **≥18 years N=297** | |
| | **Antigen+ N=27** | **Antigen– N=10** | **Antigen+ N=240** | **Antigen– N=57** |
| Rigors | 0 (0) | 0 (0) | 0 (0) | 0 (0) |
| **Days since symptom onset[b]** | | | | |
| 0–2 days since symptom onset | 15 (68.2) | 3 (42.9) | 88 (41.9) | 19 (45.2) |
| 3–5 days since symptom onset | 3 (13.6) | 3 (42.9) | 80 (38.1) | 6 (14.3) |
| 6–7 days since symptom onset | 2 (9.1) | 0 (0) | 21 (10.0) | 3 (7.1) |
| >7 days since symptom onset | 1 (4.5) | 1 (14.3) | 18 (8.6) | 11 (26.2) |
| Unknown symptom onset | 1 (4.5) | 0 (0) | 3 (1.4) | 3 (7.1) |

[a] Council of State and Territorial Epidemiologists (CSTE) clinical criteria is a surveillance case definition used within public health surveillance systems within the United States due to the non–specific nature of symptoms associated with COVID–19

[b] Percent denominator is participants reporting ≥1 symptom

**Supplementary Table 3:** Demographic information, exposure and symptoms of BinaxNOW antigen test or real–time reverse transcription–polymerase chain reaction (RT–PCR) positive participants aged <18 years by age group, Wisconsin, November–December 2020

| | RT–PCR positive no (%) | | | | | | | |
|---|---|---|---|---|---|---|---|---|
| | 5–8 years N=7 | | 9–12 years N=9 | | 13–15 years N=6 | | 16–17 years N=15 | |
| | Antigen + N=5 | Antigen– N=2 | Antigen+ N=7 | Antigen– N=2 | Antigen+ N=4 | Antigen– N=2 | Antigen+ N=11 | Antigen– N=4 |
| **Sex** | | | | | | | | |
| Male | 2 (40.0) | 1 (50.0) | 3 (42.9) | 1 (50.0) | 2 (50.0) | 2 (100) | 7 (63.6) | 1 (25.0) |
| Female | 3 (60.0) | 1 (50.0) | 4 (57.1) | 1 (50.0) | 2 (50.0) | 0 (0) | 4 (36.4) | 3 (75.0) |
| **Race/Ethnicity** | | | | | | | | |
| White, non–Hispanic | 2 (40.0) | 2 (100) | 5 (71.4) | 2 (100) | 3 (75.0) | 2 (100) | 10 (90.1) | 4 (100) |
| Hispanic/Latino | 2 (40.0) | 0 (0) | 2 (28.6) | 0 (0) | 1 (25.0) | 0 (0) | 1 (9.1) | 0 (0) |
| Unknown | 1 (20.0) | 0 (0) | 0 (0) | 0 (0) | 0 (0) | 0 (0) | 0 (0) | 0 (0) |
| **Contact with a COVID–19 case in the past 14 days** | | | | | | | | |
| Yes | 3 (60.0) | 1 (50.0) | 7 (100) | 0 (0) | 1 (25.0) | 0 (0) | 7 (63.6) | 2 (50.0) |
| Median (range) days since exposure | 9 (0–9) | 6 | 3 (0–6) | — | 3 | — | 4 (1–5) | 2 (1–2) |
| No | 0 (0) | 0 (0) | 0 (0) | 2 (100) | 2 (50.0) | 1 (50.0) | 3 (27.3) | 1 (25.0) |
| Don't know/Unknown | 2 (40.0) | 1 (50.0) | 0 (0) | 0 (0) | 1 (25.0) | 1 (50.0) | 1 (9.1) | 1 (25.0) |
| **≥1 symptom at time of testing** | | | | | | | | |
| Yes | 2 (40.0) | 0 (0) | 6 (85.7) | 1 (50.0) | 4 (100) | 2 (100) | 10 (90.9) | 4 (100) |
| No | 3 (60.0) | 2 (100) | 1 (14.3) | 1 (50.0) | 0 (0) | 0 (0) | 0 (0) | 0 (0) |
| Unknown | 0 (0) | 0 (0) | 0 (0) | 0 (0) | 0 (0) | 0 (0) | 1 (9.1) | 0 (0) |
| **CSTE clinical criteria* at time of testing** | | | | | | | | |
| Yes | 0 (0) | 0 (0) | 3 (42.9) | 0 (0) | 2 (50.0) | 1 (50.0) | 10 (90.9) | 4 (100) |
| No | | | | | | | | |
| Unknown symptom status | 0 (0) | 0 (0) | 0 (0) | 0 (0) | 0 (0) | 0 (0) | 1 (9.1) | 0 (0) |
| **Reported symptoms at time of testing**[b] | | | | | | | | |
| Congestion | 2 (100) | – | 2 (33.3) | 1 (100) | 3 (75.0) | 1 (50.0) | 7 (70.0) | 3 (75.0) |
| Sore throat | 0 (0) | – | 3 (50.0) | 0 (0) | 1 (25.0) | 0 (0) | 5 (50.0) | 1 (25.0) |
| Headache | 0 (0) | – | 0 (0) | 0 (0) | 1 (25.0) | 0 (0) | 6 (60.0) | 1 (25.0) |
| Cough | 0 (0) | – | 2 (33.3) | 0 (0) | 0 (0) | 0 (0) | 3 (30.0) | 2 (50.0) |
| Fatigue | 0 (0) | – | 0 (0) | 0 (0) | 1 (25.0) | 1 (50.0) | 3 (30.0) | 1 (25.0) |
| Muscle aches | 0 (0) | – | 0 (0) | 0 (0) | 1 (25.0) | 0 (0) | 3 (30.0) | 0 (0) |

| | **RT–PCR positive no (%)** | | | | | | | |
|---|---|---|---|---|---|---|---|---|
| | **5–8 years N=7** | | **9–12 years N=9** | | **13–15 years N=6** | | **16–17 years N=15** | |
| | **Antigen + N=5** | **Antigen– N=2** | **Antigen+ N=7** | **Antigen– N=2** | **Antigen+ N=4** | **Antigen– N=2** | **Antigen+ N=11** | **Antigen– N=4** |
| Chills | 0 (0) | – | 0 (0) | 0 (0) | 0 (0) | 0 (0) | 1 (10.0) | 0 (0) |
| Loss of smell | 0 (0) | – | 0 (0) | 0 (0) | 2 (50.0) | 1 (50.0) | 2 (20.0) | 2 (50.0) |
| Abdominal pain | 0 (0) | – | 0 (0) | 0 (0) | 0 (0) | 0 (0) | 1 (10.0) | 0 (0) |
| Nausea | 0 (0) | – | 0 (0) | 0 (0) | 0 (0) | 0 (0) | 1 (10.0) | 0 (0) |
| Fever | 0 (0) | – | 0 (0) | 0 (0) | 0 (0) | 0 (0) | 2 (20.0) | 0 (0) |
| Shortness of breath | 0 (0) | – | 0 (0) | 0 (0) | 0 (0) | 0 (0) | 3 (30.0) | 0 (0) |
| Loss of taste | 0 (0) | – | 0 (0) | 0 (0) | 1 (25.0) | 1 (50.0) | 1 (10.0) | 0 (0) |
| Diarrhea | 0 (0) | – | 0 (0) | 0 (0) | 0 (0) | 0 (0) | 1 (10.0) | 1 (25.0) |
| Rigors | 0 (0) | – | 0 (0) | 0 (0) | 0 (0) | 0 (0) | 0 (0) | 0 (0) |
| **Days since symptom onset**[b] | | | | | | | | |
| 0–2 days since symptom onset | 0 (0) | – | 4 (66.7) | 0 (0) | 3 (75.0) | 1 (50.0) | 8 (80.0) | 2 (50.0) |
| 3–5 days since symptom onset | 1 (50.0) | – | 1 (16.7) | 0 (0) | 0 (0) | 1 (50.0) | 1 (10.0) | 2 (50.0) |
| 5–7 days since symptom onset | 1 (50.0) | – | 1 (16.7) | 0 (0) | 0 (0) | 0 (0) | 0 (0) | 0 (0) |
| >7 days since symptom onset | 0 (0) | – | 0 (0) | 1 (100) | 1 (25.0) | 0 (0) | 0 (0) | 0 (0) |
| Unknown symptom onset | 0 (0) | – | 0 (0) | 0 (0) | 0 (0) | 0 (0) | 1 (10.0) | 0 (0) |

[a] Council of State and Territorial Epidemiologists (CSTE) clinical criteria is a surveillance case definition used within public health surveillance systems within the United States due to the non–specific nature of symptoms associated with COVID–19

[b] Percent denominator is participants reporting ≥1 symptom

**Supplementary Table 4:** N–gene RT–PCR mean and interquartile range (IQR) cycle threshold (Ct) values comparison by age, Oshkosh, Wisconsin, November–December 2020

| | RT–PCR Median N gene Ct (IQR) | | | | P–value |
|---|---|---|---|---|---|
| Age | **5–8 years** | **9–12 years** | **13–15 years** | **16–17 years** | |
| | 22.2 (21.4–31.6) | 22.8 (19.3–26.9) | 23.4 (15.2–29.8) | 20.6 (18.6–26.6) | 0.90 |
| **Contact with a COVID–19 case in the past 14 days** | | | | | |
| Yes | 24.4 (21.5–29.4) | 19.6 (17.8–23.2) | 23.8 | 18.8 (17.6-23.8) | 0.32 |
| No or unknown | 22.2 (16.0–31.9) | 30.7 (27.6–33.8) | 23.0 (15.2–29.8) | 24.2 (20.6–29.9) | 0.54 |
| **≥1 symptom at time of testing** | | | | | |
| Yes | 24.3 (21.4–27.3) | 19.6 (17.8–26.9) | 23.4 (15.2-29.8) | 21.6 (18.6-26.6) | 0.94 |
| No | 22.2 (21.6–31.6) | 25.4 (23.2–27.6) | — | — | 0.70 |
| **CSTE clinical criteria**[a] | | | | | |
| Yes | — | 22.8 (19.6-26.9) | 23.8 (15.1-29.8) | 21.6 (18.6-26.6) | 0.93 |
| No | 22.2 (21.4-31.6) | 21.2 (17.8-27.6) | 23.0 (15.2-32.5) | 20.21 | 0.93 |
| **Antigen test result** | | | | | |
| Positive | 21.6 (21.4–22.2) | 19.6 (17.8–23.2) | 19.1 (15.1–23.4) | 18.8 (17.6–22.5) | 0.79 |
| Negative | 31.8 (31.6-31.9) | 30.7 (27.6–33.8) | 31.2 (29.8–32.5) | 30.3 (26.9–32.2) | 0.91 |

[a] Council of State and Territorial Epidemiologists (CSTE) clinical criteria is a surveillance case definition used within public health surveillance systems within the United States due to the non–specific nature of symptoms associated with COVID–19

**Supplementary Table 5:** Sensitivity, specificity, positive predictive value, and negative predictive value of BinaxNOW antigen test compared with real–time reverse transcription–polymerase chain reaction (RT–PCR) among child and adult participants overall, by symptom status, and by exposure status, Oshkosh, Wisconsin, November–December 2020

| | RT–PCR result, no. | | | | | |
|---|---|---|---|---|---|---|
| | <18 years | | | ≥18 years | | |
| **Antigen test result** | **Positive** | **Negative** | **Total** | **Positive** | **Negative** | **Total** |
| **Overall** | | | | | | |
| Positive | 27 | 0 | 27 | 240 | 2 | 242 |
| Negative | 10 | 188 | 198 | 57 | 1586 | 1643 |
| **Total** | 37 | 188 | 225 | 297 | 1588 | 1885 |
| **Test evaluation, % (95% CI)** | | | | | | |
| Sensitivity | 73.0 (55.9–86.2) | | | 80.8 (75.9–85.1) | | |
| Specificity | 100 (98.1–100) | | | 99.9 (99.5–100) | | |
| Positive predictive value | 100 (87.2–100) | | | 99.2 (97.0–99.9) | | |
| Negative predictive value | 94.9 (90.9–97.6) | | | 96.5 (95.5–97.4) | | |
| **Symptomatic**[a] | | | | | | |
| | **Positive** | **Negative** | **Total** | **Positive** | **Negative** | **Total** |
| Positive | 22 | 0 | 22 | 210 | 1 | 211 |
| Negative | 7 | 93 | 100 | 42 | 813 | 855 |
| **Total** | 29 | 93 | 122 | 252 | 814 | 1066 |
| **Test evaluation, % (95% CI)** | | | | | | |
| Sensitivity | 75.9 (56.5–89.7) | | | 83.3 (78.1–87.7) | | |
| Specificity | 100 (96.1–100) | | | 99.9 (99.3–100) | | |
| Positive predictive value | 100 (84.6–100) | | | 99.5 (97.4–100) | | |
| Negative predictive value | 93.0 (86.1–97.1) | | | 95.1 (93.4–96.4) | | |
| **Asymptomatic**[a] | | | | | | |
| | **Positive** | **Negative** | **Total** | **Positive** | **Negative** | **Total** |
| Positive | 4 | 0 | 4 | 27 | 1 | 28 |
| Negative | 3 | 92 | 95 | 14 | 736 | 750 |
| **Total** | 7 | 92 | 99 | 41 | 737 | 778 |
| **Test evaluation, % (95% CI)** | | | | | | |
| Sensitivity | 57.1 (18.4–90.1) | | | 65.9 (49.4–79.9) | | |
| Specificity | 100 (96.1–100) | | | 99.9 (99.2–100) | | |
| Positive predictive value | 100 (39.8–100) | | | 96.4 (81.7–99.9) | | |
| Negative predictive value | 96.8 (91.0–99.3) | | | 98.1 (96.9–99.0) | | |
| **Reported contact within 14 days** | | | | | | |
| | **Positive** | **Negative** | **Total** | **Positive** | **Negative** | **Total** |
| Positive | 18 | 0 | 18 | 130 | 2 | 132 |
| Negative | 3 | 91 | 94 | 33 | 617 | 650 |
| **Total** | 21 | 91 | 112 | 163 | 619 | 782 |
| **Test evaluation, % (95% CI)** | | | | | | |

| | | | | | | |
|---|---|---|---|---|---|---|
| Sensitivity | 85.7 (63.7–97.0) | | | 79.8 (72.8–85.6) | | |
| Specificity | 100 (96.0–100) | | | 99.7 (98.8–100) | | |
| Positive predictive value | 100 (81.5–100) | | | 98.5 (94.6–99.8) | | |
| Negative predictive value | 96.8 (91.0–99.3) | | | 94.9 (92.9–96.5) | | |
| **Reported contact within 14 days and symptomatic** | | | | | | |
| | **Positive** | **Negative** | **Total** | **Positive** | **Negative** | **Total** |
| Positive | 15 | 0 | 15 | 114 | 1 | 115 |
| Negative | 2 | 30 | 32 | 24 | 298 | 322 |
| **Total** | 17 | 30 | 47 | 138 | 299 | 437 |
| **Test evaluation, % (95% CI)** | | | | | | |
| Sensitivity | 88.2 (63.6–98.5) | | | 82.6 (75.2–88.5) | | |
| Specificity | 100 (88.4–100) | | | 99.7 (98.2–100) | | |
| Positive predictive value | 100 (78.2–100) | | | 99.1 (95.3–100) | | |
| Negative predictive value | 93.8 (79.2–99.2) | | | 92.5 (89.1–95.2) | | |
| **Reported contact within 14 days and asymptomatic** | | | | | | |
| | **Positive** | **Negative** | **Total** | **Positive** | **Negative** | **Total** |
| Positive | 2 | 0 | 2 | 15 | 1 | 16 |
| Negative | 1 | 59 | 60 | 9 | 311 | 320 |
| **Total** | 3 | 59 | 62 | 24 | 312 | 336 |
| **Test evaluation, % (95% CI)** | | | | | | |
| Sensitivity | 66.7 (9.4–99.2) | | | 62.5 (40.6–81.2) | | |
| Specificity | 100 (93.9–100) | | | 99.7 (98.2–100) | | |
| Positive predictive value | 100 (15.8–100) | | | 93.8 (69.8–99.8) | | |
| Negative predictive value | 98.3 (91.1–100) | | | 97.2 (94.7–98.7) | | |

[a] Symptomatic defined as reporting ≥1 symptom at specimen collection. Asymptomatic defined as reporting no symptoms at specimen collection. Four pediatric participants and 41 adult participants with unknown symptom status not included

**Supplementary Table 6:** Sensitivity, specificity, positive predictive value, and negative predictive value of BinaxNOW antigen test compared with real–time reverse transcription–polymerase chain reaction (RT–PCR) among children by age group, overall and by symptom status, Oshkosh, Wisconsin, November–December, 2020

| | **RT–PCR result, no.** | | | | | | | | | | | |
|---|---|---|---|---|---|---|---|---|---|---|---|---|
| | **5–8 years** | | | **9–12 years** | | | **13–15 years** | | | **16–17 years** | | |
| **Antigen test result** | **Positive** | **Negative** | **Total** | **Positive** | **Negative** | **Total** | **Positive** | **Negative** | **Total** | **Positive** | **Negative** | **Total** |
| Positive | 5 | 0 | 5 | 7 | 0 | 7 | 4 | 0 | 4 | 11 | 0 | 11 |
| Negative | 2 | 35 | 37 | 2 | 53 | 55 | 2 | 60 | 62 | 4 | 40 | 44 |
| **Total** | 7 | 35 | 42 | 9 | 53 | 62 | 6 | 60 | 66 | 15 | 40 | 55 |
| **Test evaluation, % (95% CI)** | | | | | | | | | | | | |
| Sensitivity | 71.43 (29.0–96.3) | | | 77.8 (40.0–97.2) | | | 66.7 (22.3–95.7) | | | 73.3 (44.9–92.2) | | |
| Specificity | 100 (90.0–100) | | | 100 (93.3–100) | | | 100 (94.0–100) | | | 100 (91.2–100) | | |
| Positive predictive value | 100 (47.8–100) | | | 100 (59.0–100) | | | 100 (39.8–100) | | | 100 (71.5–100) | | |
| Negative predictive value | 94.6 (81.8–99.3) | | | 96.4 (87.5–99.6) | | | 96.8 (88.8–99.6) | | | 90.9 (78.3–97.5) | | |
| Symptomatic[a] | | | | | | | | | | | | |
| Positive | 2 | 0 | 2 | 6 | 0 | 6 | 4 | 0 | 4 | 10 | 0 | 10 |
| Negative | 0 | 22 | 22 | 1 | 26 | 27 | 2 | 29 | 31 | 4 | 16 | 20 |
| **Total** | 2 | 22 | 24 | 7 | 26 | 33 | 6 | 29 | 35 | 14 | 16 | 30 |
| **Test evaluation, % (95% CI)** | | | | | | | | | | | | |
| Sensitivity | 100 (15.8–100) | | | 85.7 (42.1–99.6) | | | 66.7 (22.3–95.7) | | | 71.4 (41.9–91.6) | | |
| Specificity | 100 (84.6–100) | | | 100 (86.8–100) | | | 100 (88.1–100) | | | 100 (79.4–100) | | |
| Positive predictive value | 100 (15.8–100) | | | 100 (54.1–100) | | | 100 (39.8–100) | | | 100 (69.2–100) | | |
| Negative predictive value | 100 (84.6–100) | | | 96.3 (81.0–99.9) | | | 93.5 (78.6–99.2) | | | 80.0 (56.3–94.3) | | |
| Asymptomatic[a] | | | | | | | | | | | | |
| Positive | 3 | 0 | 3 | 1 | 0 | 1 | 0 | 0 | 0 | 0 | 0 | 0 |
| Negative | 2 | 13 | 15 | 1 | 27 | 28 | 0 | 29 | 29 | 0 | 23 | 23 |
| **Total** | 5 | 13 | 18 | 2 | 27 | 29 | 0 | 29 | 29 | 0 | 23 | 23 |
| **Test evaluation, % (95% CI)** | | | | | | | | | | | | |
| Sensitivity | 60 (14.7–94.7) | | | 50 (1.3–98.7) | | | — | | | — | | |
| Specificity | 100 (75.3–100) | | | 100 (87.2–100) | | | 100 (88.1–100) | | | 100 (85.2–100) | | |
| Positive predictive value | 100 (29.4–100) | | | 100 (2.5–100) | | | — | | | — | | |
| Negative predictive value | 86.7 (59.5–98.3) | | | 96.4 (81.7–99.9) | | | 100 (88.1–100) | | | 100 (85.2–100) | | |

[a] Symptomatic defined as reporting ≥1 symptom at specimen collection. Asymptomatic defined as reporting no symptoms at specimen collection. Four pediatric participants with unknown symptom status not included

**Supplementary Table 7:** Sensitivity, specificity, positive predictive value, and negative predictive value of BinaxNOW antigen test compared with real–time reverse transcription–polymerase chain reaction (RT–PCR) among unique[a] children (n=217), Oshkosh, Wisconsin, November–December 2020

| | **RT–PCR result, no.** | | | | | | | | |
|---|---|---|---|---|---|---|---|---|---|
| | **Symptomatic** | | | **Asymptomatic** | | | **All[b]** | | |
| **Antigen test result** | **Positive** | **Negative** | **Total** | **Positive** | **Negative** | **Total** | **Positive** | **Negative** | **Total** |
| Positive | 20 | 0 | 20 | 4 | 0 | 4 | 25 | 0 | 25 |
| Negative | 7 | 89 | 96 | 3 | 90 | 93 | 10 | 182 | 192 |
| **Total** | 27 | 89 | 116 | 7 | 90 | 97 | 35 | 182 | 217 |
| **Test evaluation, % (95% CI)** | | | | | | | | | |
| Sensitivity | 74.1 (53.7–88.9) | | | 57.1 (18.4–90.1) | | | 71.4 (53.7–85.4) | | |
| Specificity | 100 (95.9–100) | | | 100 (96.0–100) | | | 100 (98.0–100) | | |
| Positive predictive value | 100 (83.2–100) | | | 100 (39.8–100) | | | 100 (86.3–100) | | |
| Negative predictive value | 92.7 (85.6–97.0) | | | 96.8 (90.9–99.3) | | | 94.8 (90.6–97.5) | | |

[a] Using the first encounter for repeat participants

[b] Includes 4 participants with unknown symptom status
